# SERPINA12 Splice Site Variant in Tunisian Familial Psoriasis Vulgaris: Segregation to Inheritance

**DOI:** 10.64898/2026.09.11.26362406

**Authors:** Mariem Ennouri, Slaheddine Marrakchi, Fakher Frikha, Emna Bahloul, Anouar Bel Hadj Ltaief, Rania Ammar, Hamida Turki, Noura Bougacha Elleuch

**Affiliations:** Laboratory of Molecular and Functional Genetics, Faculty of Sciences of Sfax, Sfax University, Sfax 3029, Tunisia; Department of Dermatology, CHU Hedi Chaker, Sfax University, Sfax 3029, Tunisia; Laboratory of Molecular and Cellular Screening Processes, Centre of Biotechnology of Sfax, University of Sfax, P O Box 1177, Sidi Mansour Road, 3018, Sfax, Tunisia; Primary healthcare group Sfax, TUNISIA

**Keywords:** Psoriasis vulgaris, T2DM, SERPINA12, KLK7, Vaspin

## Abstract

**Background:** Psoriasis vulgaris (PV) is an inflammatory skin disorder whose genetic basis has been widely explored through genome-wide association studies and biomarker analyses in heterogeneous cohorts. We adopted a family-based strategy focusing on nuclear families.

**Objective:** To identify genetic variants associated with PV and elucidate their impact on disease pathogenesis and metabolic comorbidities.

**Methods:** Whole exome sequencing (WES) was performed on a PV affected nuclear families. Integrative computational analyses, including structural modeling and molecular dynamics simulations, were used to evaluate the impact of identified variants on protein stability and function. Potential links to inflammation and metabolic dysfunction were explored through literature-based interpretation.

**Results:** We identified a heterozygous splice site variant c.905+1 G>T in the SERPINA12 gene that segregates with PV in the studied families. Structural modeling and molecular dynamics revealed destabilization of the C-terminal region, impairing its serine protease inhibitory function. This alteration reduces vaspin’s anti-inflammatory activity via kallikrein 7 (KLK7) inhibition.

**Conclusion:** This integrative WES-based approach identifies a PV associated mutation and suggests a new pathogenic mechanism involving the vaspin/KLK7 axis. As these molecules are expressed in the skin and may influence insulin activity, these findings offer insight into the link between PV and type 2 diabetes mellitus and highlight potential targets for precision medicine in psoriasis and associated metabolic disorders.

## INTRODUCTION

Psoriasis vulgaris (PV) is a chronic inflammatory skin disease affecting approximately 2–3% of the global population.^1^ Increasing evidence indicates that PV is not merely a skin disorder but an auto-inflammatory disease associated with several comorbidities, including psoriatic arthritis, inflammatory bowel disease, and type 2 diabetes mellitus (T2DM).^2–4^

Clinically, PV is characterized by recurrent erythematous and scaly plaques resulting from keratinocyte hyperproliferation and dysregulated immune responses. The disease arises from a complex interplay between genetic susceptibility and environmental factors, including infections, psychological stress, hormonal changes, and medications. The substantial contribution of genetic factors is supported by twin studies demonstrating significantly higher concordance rates in monozygotic compared with dizygotic twins.^5^

The mode of inheritance of PV has long been debated. Since the 1960s and 1970s ^6^, several models have been proposed, including autosomal dominant with incomplete penetrance. While genome-wide association studies (GWAS) have demonstrated the involvement of numerous susceptibility genes^7^, no specific genetic defects were reported to be consistently responsible for PV development. Consequently, alternative approaches are needed to better characterize the genetic architecture of PV and ultimately enable precision medicine strategies.

In contrast to generalized pustular psoriasis (GPP), a rare psoriasis subtype that follows an autosomal recessive inheritance pattern and is more prevalent in consanguineous populations^8^, PV is a common disease in non-consanguineous populations. Such epidemiological observations raise the possibility that PV may follow an autosomal dominant mode of inheritance, potentially with incomplete penetrance. Similar inheritance patterns have been described in monogenic forms of diabetes, such as maturity-onset diabetes of the young (MODY)^9-11^, where causal genes were successfully identified through the study of nuclear families.

To minimize genetic heterogeneity and increase the power to detect disease associated variants, we focused on carefully selected nuclear families with multiple affected individuals. We adopted a family based Whole exome Sequencing (WES) strategy facilitate the identification of potentially pathogenic variants.

## METHODS

### i. SELECTION OF PARTICIPANTS

Eight PV patients from 3 nuclear consanguineous families from the same region in Tunisia (Figure 1) (Figure available upon request) were studied. Genetic analysis proceeded in two steps. First, whole exome sequencing (WES) was performed in three characterized patients and one aged healthy control relative, based on the expected genetic homogeneity of the cohort. Second, Sanger sequencing was performed to check segregation of the candidate variant in 5 additional affected relatives, 10 healthy relatives (Table 1) and 51 unrelated healthy Tunisian persons. Given the association between PV and T2DM, diabetes history was collected for all participants.

**Table 1:** Clinical features of psoriatic patients.

| ID | Clinical status | Sex | Age group (years) | SERPINA12 variation | T2DM |
| --- | --- | --- | --- | --- | --- |
| A.II.1 | Multiple PV lesions | F | 66-70 | +/- | No |
| B.II.2 | Non-psoriatic | M | 76-80 | +/- | Yes |
| B.II.4 | Non-psoriatic | F | 81-85 | +/- | Yes |
| B.II.5 | Non-psoriatic | M | 86-90 | -/- | No |
| B.III.1 | Non-psoriatic | F | 46-50 | -/- | No |
| B.III.2 | Non-psoriatic | F | 51-55 | +/- | No |
| B.III.4 | Multiple PV lesions | M | 51-55 | +/- | Yes |
| B.III.5 | Scrotal tongue | F | 46-50 | +/- | No |
| B.III.6 | Non-psoriatic | M | 46-50 | +/- | No |
| B.IV.1 | Multiple PV lesions | M | 21-25 | +/+ | No |
| C.I.1 | Non-psoriatic | F | 96-100 | -/- | No |
| C.II.1 | Non-psoriatic | F | 71-75 | -/- | Yes |
| C.II.2 | Non-psoriatic | M | 76-80 | +/- | Yes |
| C.II.3 | Multiple PV lesions | F | 71-75 | +/- | No |
| C.II.4 | Non-psoriatic | M | 81-85 | -/- | Yes |
| C.III.1 | Multiple PV lesions | F | 51-55 | +/- | Yes |
| C.III.2 | Multiple PV lesions | M | 46-50 | -/- | No |
| C.III.3 | Non-psoriatic | F | 46-50 | -/- | No |
| C.IV.1 | lower limbs | F | 26-30 | +/- | No |

+/-: homozygous state, +/- : heterozygous state, -/-: absent, 0: unaffected, 1: affected,
M: male, F: female, a: patient with PV but who did not carry the variation, +: confirmed associated disease, -: no associated disease at the diagnosis of PV. T2DM: type2 diabetes mellitus.

### ii. DATA COLLECTION AND MEASUREMENTS

#### DNA extraction and WES

Genomic DNA was extracted from peripheral blood using the standard phenol-chloroform method. All samples were submitted along with a requisition form that included signed informed consent and detailed clinical information. WES was performed in three affected individuals (A.II.1, B.III.4, C.II.3) and one unaffected relative (C.I.1) (Figure 1) (Figure available upon request) using the SureSelect Human All Exon 50 Mb enrichment kit (Agilent) on an Illumina HiSeq platform. Variants were prioritized according to a minor allele frequency (MAF) ≤1% in gnomAD, predicted high functional impact, and dominant inheritance

#### Sanger Sequencing

Candidate causative variant was subsequently validated and segregation assessed by PCR and sanger sequencing in affected (5) and unaffected (61): 10 healthy members in the family and 51 healthy from the general population) individuals using the primers (F:5’TGTAGGCAATGGGGTTATCA3’; R:5’TTCTTTCCACACCAACATGC3’). PCR consisted of an initial denaturation at 94°C for 2 min, followed by 32 touchdown cycles (65– 55°C) and a final extension at 72°C for 10 min

#### In silico analysis

Splice site prediction was performed using the Fruitfly prediction tool (https://fruitfly.org) for both wild-type and mutated sequences.

#### Protein Modeling

Wild-type (residues 37–414) and truncated (residues 37–302) vaspin structures were generated by homology modeling in SPDBV using the human vaspin crystal structure (PDB ID: 4IF8). Energy minimization was performed in GROMACS with the CHARMM36m force field.

#### Molecular Dynamics Simulations

Wild-type and truncated vaspin models were subjected to 100 ns molecular dynamics simulations in GROMACS 2025.0 under physiological conditions. Systems were solvated with TIP3P water, neutralized, and equilibrated under NVT and NPT conditions before production runs. Structural stability was assessed using root-mean-square deviation (RMSD), root-mean-square fluctuation (RMSF), and radius of gyration (Rg).

## RESULTS

### Molecular investigation

#### WES

We considered the disease as autosomal dominant in the analysis of WES. The *SERPINA12* located on chromosome 14 emerged as the most likely contributor to PV, given its established involvement in skin biology and functional relevance. SERPINA12 encodes for the serine protease inhibitor, vaspin. Although two additional genes, *SLC9B1* and *FRG1*, were identified in the analysis, they were excluded from further investigation due to the lack of direct biological relevance to the present study.

The c.905+1G>T variant located in the intronic region of *SERPINA12* was identified in heterozygous state in the three investigated patients. It affects a splice donor site with a high impact of severity. *In silico* prediction tools suggest that this variant is likely pathogenic (CADD score: 33.0; predicted loss of function [pLoF], high confidence). According to gnomAD, this variant has an allele frequency of 1.08 e□□, with no homozygous individuals observed.

### Clinical history of PV and T2DM

The family members were followed until 2025. In total 19 individuals were genetically analyzed. Eight were affected by PV and 11 remained unaffected. Among affected individuals, six experienced at least one episode of generalized PV, one presented with scrotal tongue, and one with localized lower-limb PV (Table 1).

Seven individuals developed T2DM whereas twelve remained diabetes-free during follow-up.

### Variant Segregation analysis (Table 1)

Among the 19 individuals included in the segregation analysis, 12 carried the SERPINA12 variant (11 heterozygous and 1 homozygous), while 7 were non carriers. Among the eight PV patients, seven carried the variant (87.5%), whereas one affected individual lacked it (12.5%). Among the eleven unaffected relatives, five were heterozygous carriers (45.45%) and six were non carriers (54.54%). The variant was absent in 51 unrelated healthy Tunisian controls. Overall, 7 of 12 variant carriers were affected by PV (58.33%), compared with 1 of 7 non-carriers (14.3%).

### Association between *SERPINA12* variation and T2DM (Table1)

Of the 12 carriers of the SERPINA12 variant, 5 developed T2DM and 7 did not. Among the 7 non-carriers, T2DM was observed in 2 individuals.

### Bioinformatic prediction

According to the Ensembl database, the *SERPINA12* gene (transcript ENST00000341228) consists of six exons and six introns. The analysis focused on exon 4 and the flanking intronic region between introns 4 and 5, which harbors the identified mutation. Splice site prediction analysis indicated that the mutation is located at the donor splice site within intron 4-5. This mutation disrupts the original donor site at position 205-219 and creates a novel donor site at position 210-224 (Table 2). Notably, no effect was observed on the acceptor splice sites. The shift in the donor position leads to the retention of five intronic nucleotides (TTAAA), which results in the introduction of a premature stop codon replacing a 303-valine residue (Figure S2).

**Table 2:**
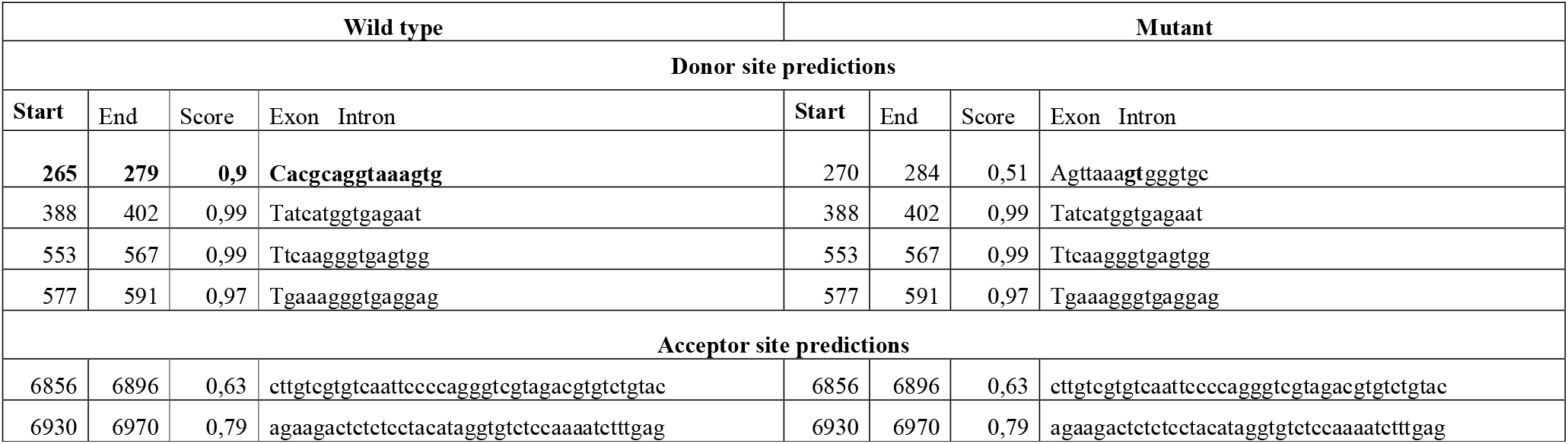
Effect of the mutation on splice site prediction using the Fruitfly tool.

As shown in Figure S1, the 3D structure of the protein highlights the deleterious region that will be absent due to the presence of the mutation, indicated in red. The remaining conserved region, shown in green, appears structurally unstable.

### Molecular modeling and molecular dynamics simulations

Molecular dynamics simulations revealed marked structural destabilization of truncated vaspin (Figure S3). Wild-type vaspin rapidly reached a stable RMSD (0.22–0.25 nm), whereas the truncated protein showed progressive deviation (up to 0.7 nm). Consistently, the radius of gyration decreased from 2.16 to 2.05 nm in the truncated model, indicating partial structural collapse, while remaining stable in the wild type (2.20–2.22 nm). RMSF analysis further demonstrated increased flexibility throughout residues 50–300 in the truncated protein compared with the wild type. Together, these findings indicate that C-terminal truncation markedly compromises vaspin structural stability and is likely to impair its biological function

## DISCUSSION

The genetic contribution to PV is widely recognized, although disease onset and severity are influenced by environmental triggers^12-14^. Despite substantial evidence supporting a hereditary component, the mode of inheritance remains incompletely understood. Early epidemiological investigations based on familial aggregation and pedigree analyses suggested an autosomal dominant pattern with incomplete penetrance^6^. More recently, GWAS in large and ethnically diverse cohorts have identified numerous susceptibility loci associated with PV improving disease understanding but not allowing identification of specific causal genes. To overcome these limitations, we adopted a family-based WES approach in consanguineous and endogamous families.

This new approach allowed the identification of a pathogenic variant, c.905+1G>T, in the SERPINA12 gene in several consanguineous and endogamous families. The variant was detected in all affected individuals from families showing segregation of the mutation (families A, B, and C). Among the 8 affected individuals, six patients carried the mutation in the heterozygous state, supporting the previously proposed autosomal dominant mode of inheritance of PV^6^. One patient was homozygous for the variant and developed the disease at an earlier age than the heterozygous carriers, suggesting a potential gene dosage effect on disease onset. Ashner raised this assumption in patients with generalized PV^15^. The mutation was absent in one affected individual whose mother belonged to a different family in which no segregation of the variant was observed. This finding suggests limited genetic heterogeneity within the studied pedigrees. This hypothesis is supported by the observation that several affected members of his maternal family developed PV despite the absence of the SERPINA12 variant (data not shown).

Furthermore, the variant was not detected in any of the 51 healthy controls analyzed, providing additional evidence supporting its pathogenic role in PV.

Among 12 carriers of the SERPINA12 variation, 7 were diagnosed with psoriasis (58.33%), giving a risk to develop the disease in carriers, close to the estimation reported by Abele^16^ who considered PV as an autosomal dominant disease with incomplete penetrance evaluated at 60%. Also, in the two nuclear families (B and C), some parents of affected children carried the mutation but remained asymptomatic (example: B.III.2, C.II.2; Figure1) (Figure available upon request), suggesting incomplete penetrance of the disease phenotype. In a cohort of 815 individuals from the same region of the United States, a high incidence of PV was reported, however, not all affected children had an affected parent, suggesting the existence of asymptomatic carriers transmitting the disease-associated allele^16^. So, when studying these common autosomal dominant inflammatory diseases like PV and T2DM by WES, we suggest to consider at least one of the parents to be genetically carrier of the potential variation in case of both parents do not express the disease.

Among the unaffected family members, the variant was absent in six individuals and present in the heterozygous state in five individuals. These asymptomatic carriers may represent presymptomatic individuals or carriers who may never develop the disease due to incomplete penetrance and the multifactorial nature of PV^17^. Therefore, the presence of the SERPINA12 variant alone may not be sufficient to trigger disease development, and additional epigenetic, or environmental factors are likely required for phenotypic expression.

Furthermore, the variant was not detected in any of the 51 healthy controls analyzed, providing additional evidence supporting its pathogenic role in PV.

The SERPINA12 gene encodes vaspin, a serine protease inhibitor originally identified in visceral adipose tissue. Vaspin is now recognized as an adipokine with significant expression in skin tissue^18^, and recent studies have suspected its involvement in pathogenesis of PV^19^ and palmoplantar keratoderma (PPK)^20^. Mohamad et al ^20^ reported two homozygous patients presenting early onset recessive form of PPK.

Vaspin is a member of the serpin family and functions as an inhibitor of KLK related peptidases, particularly KLK7^21^. It was demonstrated that transgenic mice overexpressing KLK7 develop psoriasis skin lesions^22,23^ and KLK7 expression and activity in psoriatic lesions increased in comparison with healthy and non-lesional skin in individuals with PV^24^. According to Saalbach^21^, vaspin is closely linked to keratinocyte differentiation. Lesional psoriatic skin showed a marked reduction of vaspin expression compared to normal skin in psoriatic patients and healthy skin. Downregulation of vaspin was associated with increased expression of interferon inducible genes and proinflammatory cytokines^21^. Also, in cell culture release of the proinflammatory cytokines TNFα and IL-6 as well as chemokine IL-8 were significantly inhibited by vaspin but not by a direct mechanism^21^. In contrast IL-12 and IL-23, major pathways in PV were not affected^21^ demonstrating the diversity of mechanisms in the disease and the probable pathways leading to the same clinical aspect. KLK7 could play a key role in psoriasis by promoting inflammation through pro-IL-1β activation, which triggers both local and systemic immune amplification^25^.

The identified c.905+1G>T variant likely destabilizes vaspin protein or impairs its expression, resulting in reduced inhibition of KLKs and, consequently, increased epidermal inflammation by promoting proinflammatory cytokines.

It is well known that PV is frequently associated with metabolic syndrome, a cluster of conditions including obesity, hypertension, and T2DM^26^. This association suggests common pathogenic mechanisms. Furthermore, clinical observations have shown improvement of psoriasis symptoms in diabetic patients treated with exenatide, a GLP-1 receptor agonist, suggesting that shared molecular or genetic pathways may link both diseases^27^.

Vaspin activity has been reported to be related to T2DM and metabolic syndrome^28,29^. Protective activity of vaspin is expressed by its inhibitory effect on human insulin degradation by KLK7^25^. Thus, vaspin promotes glucose tolerance and mitigates metabolic inflammation. Through KLK7 inhibition in adipose tissue, vaspin may protect against insulin degradation and support glucose homeostasis. Ribas-Latre^30^ demonstrated that KLK7 expression in adipose tissue is associated with inflammation and insulin resistance in high fat diet fed mice, reinforcing the biological relevance of the vaspin-KLK7 axis in systemic metabolic regulation. It was also found in animal model that KLK7 knockout had similar effects on metabolic health in diet induced obese mice as overexpression of vaspin^31^. Then we may hypothesize that the loss of function caused by the described variant (c.905+1G>T) may compromise the protective effects of vaspin, potentially predisposing individuals not only to psoriasis but also to insulin resistance.

In the studied families, a significant number of PV patients developed T2DM, mainly individuals over 50 years of age. Younger patients with PV did not develop metabolic syndrome, confirming the multifactorial nature of these inflammatory diseases^32^.

As other suggested to target KLK7 to treat metabolic syndrome, obesity and T2DM, we think that targeting KLK7 should be considered in the treatment of diseases induced by genetic abnormalities of SERPINA12, like PV or metabolic syndrome^30,31^.

## Conclusion

For years, genetic basis of PV has been studied mainly by GWAS and by investigating the biomarkers of the disease in large and heterogeneous populations. However, with the aim to develop targeted therapies for each patient with inflammatory disease, these approaches showed some limitation regarding the global approach of large and heterogeneous populations. Our approach of studying nuclear families allows to lower the risk to have several causative genes in large groups of unrelated patients, that could hamper the genetic analysis. This study identifies for the first time a *SERPINA12* splice site variant associated with familial psoriasis with a dominant mode of transmission with incomplete penetrance and provides a mechanistic explanation for its pathogenicity through structural modeling and molecular dynamics simulations.

These insights strengthen the rationale for targeting the vaspin-KLK axis in therapeutic strategies for psoriasis and related metabolic disorders.

## Data Availability

Whole exome sequencing data supporting the findings of this study are available from the corresponding author upon reasonable request, subject to ethical approval and data protection regulations. Structural modeling data and molecular dynamics simulation results are available

## ACKNOWLEDGEMENTS

We gratefully thank all participating family members for their valuable cooperation and for providing blood samples. We also thank all members of the Department of Dermatology, University Hospital of Sfax, Tunisia, for their support. This study was funded by the Ministry of Higher Education and Scientific Research of Tunisia (LR16E16). We acknowledge Prof. Judith Fischer and Prof. Andreas Zimmer from the Institute of Human Genetics, Medical Center University of Freiburg for their scientific support and collaboration

