## Supplemental figure 3 for "SERPINA12 Splice Site Variant in Tunisian Familial Psoriasis Vulgaris: Segregation to Inheritance"

| a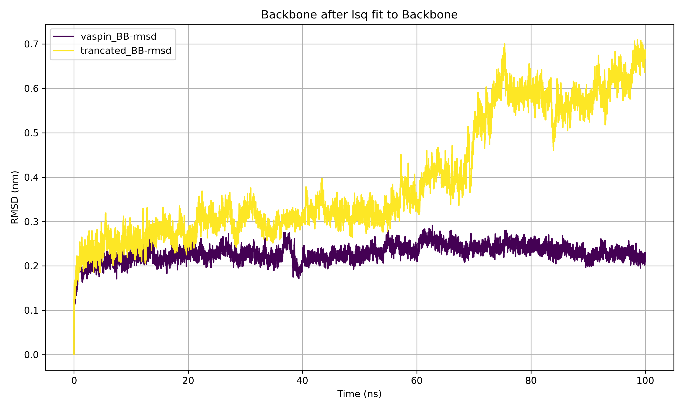 | b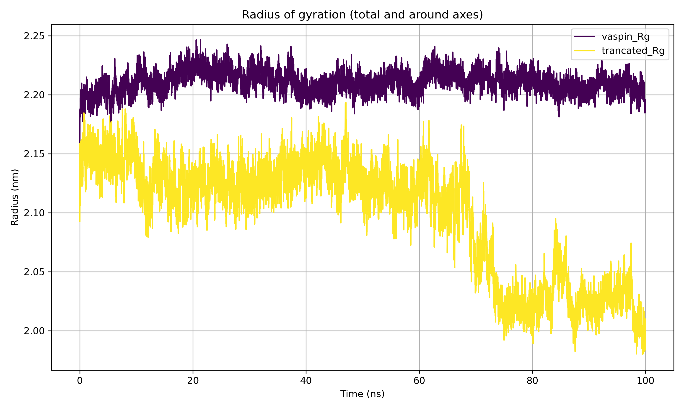 |
| --- | --- |
| c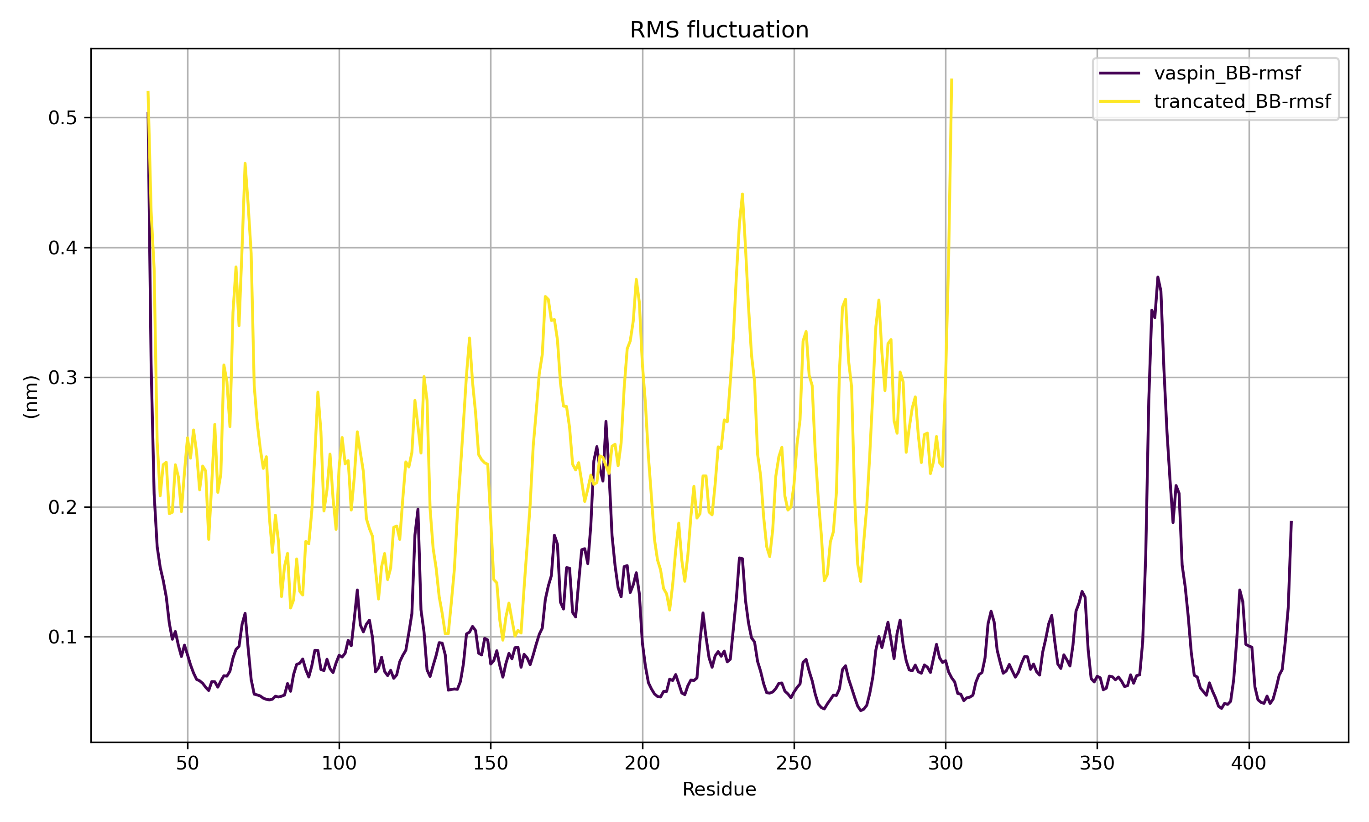 | |

**Fig.3.S: Comparative molecular dynamics analysis of wild-type and truncated vaspin proteins** (a,b,c).

(a) The Root Mean Square Deviation (RMSD, in nm) reflects global structural stability over 100 ns; the truncated protein shows a continuous increase, indicating progressive destabilization, whereas the native structure remains stable. (b) The radius of gyration (Rg, in nm) assesses structural compactness, revealing stable dimensions in the native form and a decreasing trend in the truncated variant, suggesting collapse. (c) The Root Mean Square Fluctuation (RMSF, in nm) per residue indicates local flexibility, with the truncated form showing higher internal fluctuations compared to the native protein, consistent with a loss of intramolecular cohesion
