## Supplemental figure 1 for "SERPINA12 Splice Site Variant in Tunisian Familial Psoriasis Vulgaris: Segregation to Inheritance"

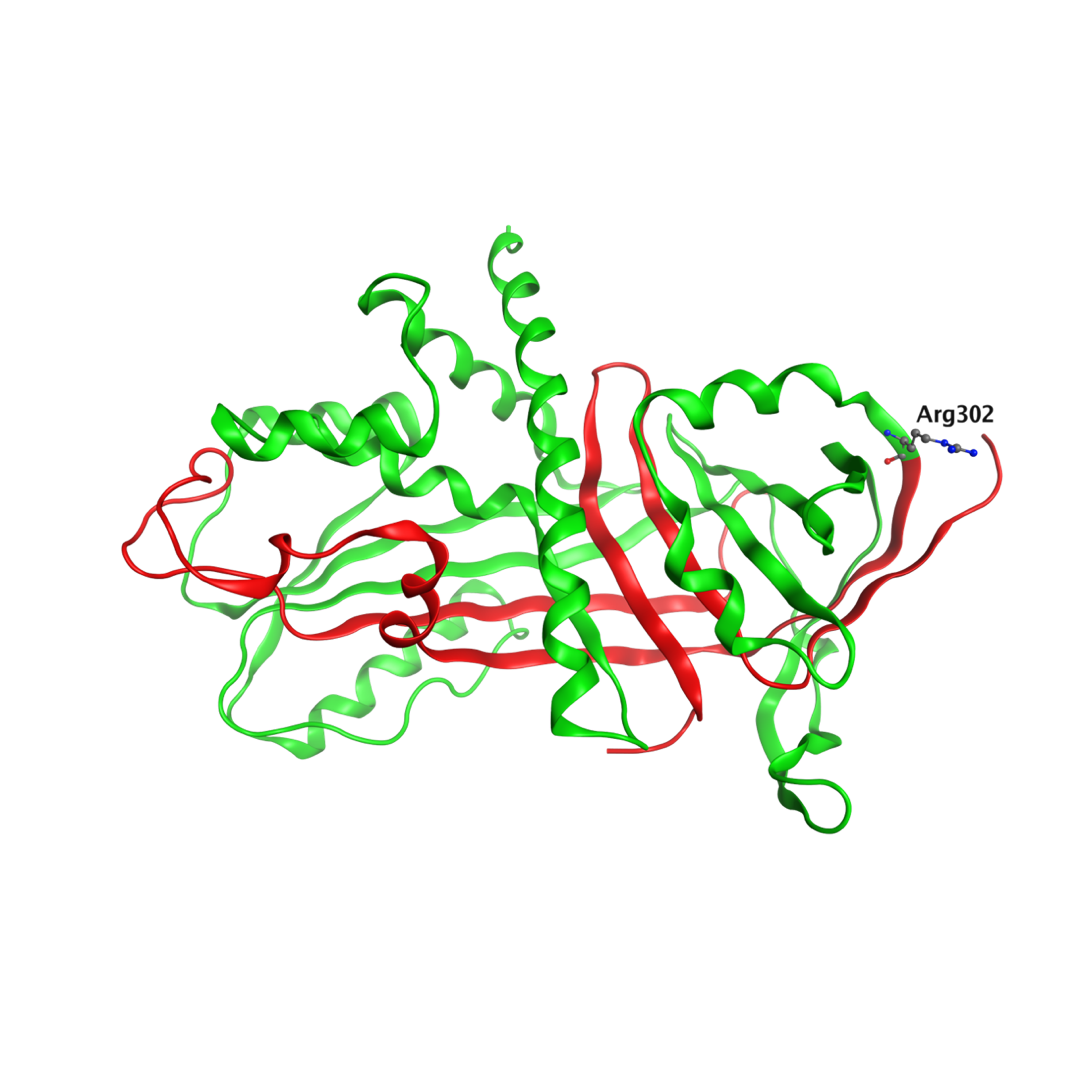

**Fig.1.S: 3D cartoon representation of the vaspin protein structure**

The conserved region is shown in green, while the red region represents the portion predicted to be absent due to the mutation. The native form (purple) and the truncated form (yellow) were compared based on three key parameters.
