## Supplemental figure 2 for "SERPINA12 Splice Site Variant in Tunisian Familial Psoriasis Vulgaris: Segregation to Inheritance"

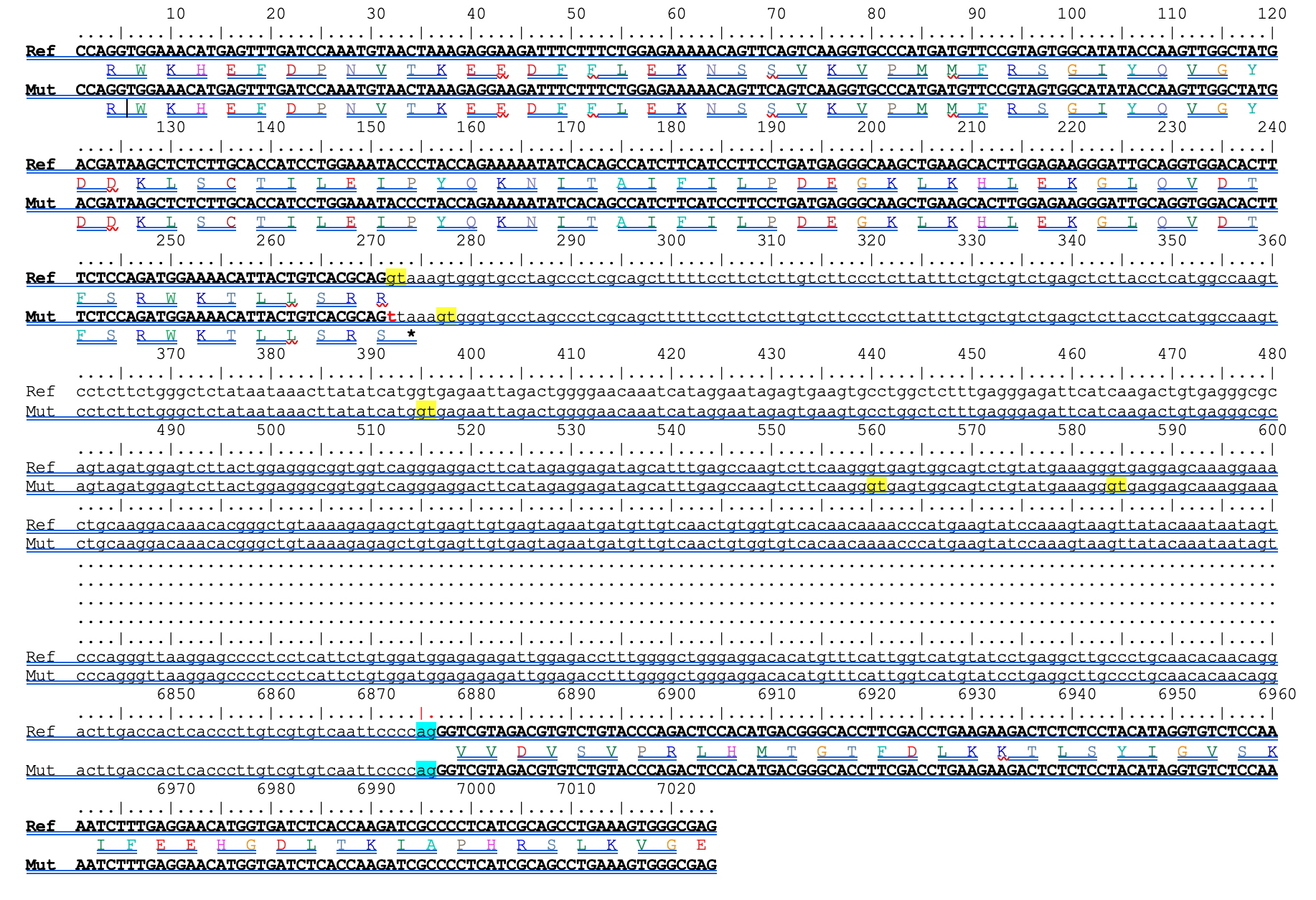

**Fig.2.S: Alignment of the wild-type (Ref) and mutated (Mut) sequences.** Donor splice sites are highlighted in yellow, acceptor sites in blue, and the mutation is indicated in red. The asterisk (*) marks the location of the premature stop codon. Splice site predictions were obtained using the Fruitfly tool.
